# Intention to receive a COVID-19 vaccine by HIV status among a population-based sample of women and gender diverse individuals in British Columbia, Canada

**DOI:** 10.1101/2021.08.19.21262249

**Authors:** Angela Kaida, Lori A. Brotto, Melanie C.M. Murray, Hélène C. F. Côté, Arianne Y. Albert, Valerie Nicholson, Rebecca Gormley, Shanlea Gordon, Amy Booth, Laurie W. Smith, Ally Baaske, Liisa A. M. Galea, Manish Sadarangani, Gina S. Ogilvie

## Abstract

**Introduction:** COVID-19 vaccination is recommended for people living with HIV (PLWH), among whom social inequities and co-morbidities may drive risks of COVID-19 infection and outcome severity. Among a provincial (British Columbia) sample, we determined the prevalence of COVID-19 vaccine intention by HIV status and assessed socio-demographic, vaccine hesitancy, and psychological predictors of vaccine intention.

**Methods:** Individuals (25-69y) recruited from province-wide research cohorts completed an online survey examining COVID-19 impacts (August/2020-March/2021). Among women and gender diverse participants, we compared intention to receive a recommended COVID-19 vaccine (Very likely/Likely vs Neutral/Unlikely/Very Unlikely) by self-reported HIV status. Logistic regression models assessed the independent effect of HIV status and other factors on vaccine intention.

**Results:** Of 5,588 participants, 69 (1.2%) were PLWH, of whom 79.7% were on antiretroviral therapy. Intention to vaccinate was significantly lower among PLWH compared to participants not living with HIV (65.2% vs 79.6%; OR: 0.44; 95%CI: 0.32-0.60). However, this association was attenuated after adjustment for social disparities (aOR:0.85; 95%CI: 0.48-1.55). Among PLWH, those with greater vaccine confidence, positive attitudes towards the COVID-19 vaccine, and more strongly influenced by direct and indirect social norms to vaccinate had significantly higher odds of vaccine intention.

**Discussion:** Tailored messaging is needed to build vaccine confidence, address questions about vaccine benefits, and support informed vaccination decision-making to promote COVID-19 vaccine uptake among women and gender diverse PLWH.

## INTRODUCTION

The COVID-19 pandemic and the associated public health response has significantly disrupted lives and livelihoods in Canada and around the world. As of July 26^th^ 2021 in Canada, 1,427,342 COVID-19 cases and 26,553 related deaths have been reported. Sex-disaggregated data reveal that 50.3% of COVID-19 cases and 49.8% of deaths are among females (1), with disproportionate impacts among individuals and communities confronting socio-structural inequities (2,3).

Early in the pandemic, the US Centres for Disease Control and Prevention flagged that people living with HIV (PLWH) may be at heightened risk of severe COVID-19 illness (4). Accumulating data suggest, however, that HIV infection itself does not confer higher susceptibility to COVID-19, (5-7) rather, HIV-accompanying social disparities and co-morbidities may drive observed increases in the risk of infection and outcome severity among PLWH (8,9). This distinction is important as it informs government and public health officials on how to best act to reduce inequities.

The National Advisory Committee on Immunization (NACI) in Canada considered such social disparities and co-morbidities, alongside considerations of risks for SARS-CoV-2 infection and severe illness, to identify priority populations for the first phase of COVID-19 vaccination (10). Canada launched its COVID-19 vaccine roll-out in December 2020 for adults, with eligibility expanding to include all individuals 12+ years of age (without contraindications) by June 2021 (11).

Although relatively few PLWH participated in COVID-19 vaccine trials, available data indicate that the vaccines are effective and that there are no unusual safety concerns among people with well-controlled HIV, including those with undetectable viral loads and CD4 cell counts above 200 cells/mm^3^ (12,13). As such, the NACI strongly recommended that immunosuppressed and immunocompromised individuals (including PLWH) be offered a complete COVID-19 vaccine series (10). In tandem, the British Columbia Centre for Excellence in HIV/AIDS (BCCfE) Committee for Drug Evaluation and Therapy similarly advised that “*People living with HIV (PLWH) aged 18 years or older should be vaccinated for COVID-19 if they meet current public health criteria for priority groups and if they have no contraindications… regardless of CD4 count*”, and recommends receipt of any of the COVID-19 vaccines currently approved in Canada (i.e., Pfizer-BioNTech, Moderna, AstraZeneca, and Janssen vaccines) (14).

Adherence to these recommendations and the ultimate success of the national COVID-19 vaccine roll-out is contingent on vaccine intention and vaccine uptake. Vaccine hesitancy (a concept defined as the refusal or delay in accepting vaccination despite the availability of vaccination services (15,16)), vaccine misinformation, and medical mistrust may limit vaccine uptake and contribute to further perpetuating COVID-19 inequities (17-19). There are currently few data regarding intention to receive the COVID-19 vaccine among PLWH (17), and, to our knowledge, no data from women or gender diverse individuals living with HIV. Moreover, there is a paucity of data examining vaccine hesitancy or the attitudes, social norms, and perceived behavioral controls that predict COVID-19 vaccine intention among PLWH, and whether these differ from patterns in the general population. In Canada, such data are particularly pertinent since women living with HIV (WLWH) experience significant socio-structural inequities and co-morbidities relative to both men living with HIV and HIV-negative women. For instance, among WLWH, 79% are Indigenous, Black, or other women of colour, including 36% who are of Indigenous ancestry (20); 70% live below the poverty line ($20K CAD per year) (21); and 75% live with one or more co-morbidities in additional to HIV, including cardiovascular disease, cancers, osteoporosis, chronic kidney or liver disease, chronic depression, anxiety and other mental health illnesses (22,23). WLWH also have poorer HIV clinical outcomes across the HIV care cascade including lower prevalence of antiretroviral therapy (ART) initiation and HIV viral suppression compared with men (24). All factors known to increase risk and consequence of SARS-CoV-2 infection.

Using population-based survey data from a provincial sample of women and gender diverse individuals in British Columbia (BC), Canada, the objectives of this study were (1) to estimate and compare intention to receive the COVID-19 vaccine by HIV status; (2) to measure and compare the prevalence of vaccine hesitancy (16,25) by HIV status; (3) to measure and compare the prevalence of four COVID-19 vaccine-specific psychological constructs grounded in the Theory of Planned Behavior (26) by HIV status, including vaccine attitudes, perceived behavioral control to receive the COVID-19 vaccine if desired, and the influence of direct and indirect social norms; and (4) among those living with HIV, to examine whether vaccine hesitancy and psychological constructs predict COVID-19 vaccine intention.

These analyses are aimed at guiding public health programming and recommendations for COVID-19 vaccination for women and gender diverse individuals living with HIV to optimize COVID-19 vaccine uptake in this population.

## METHODS

### Study design and participants

We used cross-sectional survey data from participants enrolled in the **R**apid **E**vidence **S**tudy of a **P**rovincial **P**opulation Based C**O**hort for Ge**N**der and **SE**x (RESPPONSE) study, which assessed the impacts of COVID-19 and the associated public health control measures on people across the Canadian province of BC (27).

Individuals (aged 25-69 years, BC residents) enrolled in existing, large provincially-representative community and hospital-based cohort studies who had consented to be contacted for future research were invited to complete an online survey examining impacts of COVID-19 (August 20-March 1, 2021) and receive an at-home SARS-CoV-2 research antibody test (results to be reported elsewhere). Two existing cohorts (the Canadian HIV Women’s Sexual and Reproductive Health cohort study (CHIWOS) (28) and the Children and Women: AntiRetroviral Therapy and Markers of Aging (CARMA) study (22) specifically enrolled WLWH while other cohorts enrolled members of the general population, inclusive to all people living with HIV.

All eligible individuals were sent an email invitation to participate in an online survey. To increase sex and gender diversity of the study, upon completing the survey, participants were asked to provide the email address of an adult household member who identified as another gender. These individuals were then invited to participate. All prospective participants who did not complete the survey after the initial invitation were sent up to two email reminders, each seven days apart. Participants who did not complete the survey within 21 days after the initial invitation were considered as having declined participation.

For power considerations, we aimed to enroll a total of n=750 participants per each 5-year age-strata (19). After recruiting from the existing cohorts, we pursued public recruitment via social media, websites, listservs, and word-of-mouth to fill the target quota for individuals aged 25-40 and 65-69 years. We employed additional targeted recruitment strategies to enhance study participation among WLWH (of all eligible ages), who are consistently under-represented in research (29,30). Learning from community-based research principles (31),(32), we hired and trained three experienced Peer Research Associates (WLWH trained in quantitative research methods) (33) to support recruitment of WLWH, who may not have had a working email address, reliable access to computers, internet access, or other infrastructure required to complete an online survey. We also pursued recruitment of WLWH via researchers, HIV clinics, and community-based organizations who support PLWH in BC.

### Ethical Considerations

All participants provided voluntary informed consent at enrollment. After completing the survey, participants were entered into a lottery to receive a $100 gift card. Ethical approval was received from The University of British Columbia Research Ethics Board (H20-01421).

### Inclusion and exclusion criteria

Analyses were restricted to self-identified women and gender diverse participants either living with or not living with HIV. Gender diverse individuals comprised 1.2% of the overall sample (19), however, given a high proportion of gender diverse individuals living with HIV who identified a biological sex of female, we chose to include this group in the analysis to enable consideration of this priority and underserved group living with HIV.

### Study Procedures

Participants completed a structured online questionnaire (supported by Research Electronic Data Capture (REDCap)) software (34). The questionnaire was developed by experts in sex-and-gender based analysis, vaccine intention, Theory of Planned Behavior, social determinants of health, economics, mental health, and sexual and reproductive health, using validated scales when available. The questionnaire was assessed for face validity and comprehension, pilot tested, revised, and the final version was implemented using REDCap. Questionnaires were available in English and took a median of 31 minutes [Interquartile range [IQR]: 23-47] to complete.

### Measures

The primary outcome was ‘intention to vaccinate’, considered as the most proximate measure to actual vaccine uptake, and assessed via a 5-point Likert scale to the question “*If a COVID-19 vaccine were to become available to the public, and recommended for you, how likely are you to receive it*?” The question was phrased theoretically given that a large majority of participants completed the survey before the COVID-19 vaccine was widely available in BC. Consistent with a RESPPONSE study analysis of overall vaccine intentions in BC, responses were dichotomized as follows: Participants who reported “Very Likely” or “Somewhat Likely” were considered as having an intention to vaccinate while those who reported “Neutral”, “Unlikely”, or “Very Unlikely” were considered as not intending to vaccinate (19).

Potential socio-demographic correlates of vaccine intention were considered *a priori*, including: age, sex, gender (woman or gender diverse, which referred to individuals who identify as, but not limited to, gender non-binary, GenderQueer, Two-Spirit, agender, gender fluid, gender non-conforming, or other gender identity), Indigenous ancestry, ethnicity (35), education, annual household income, existing chronic health conditions (excluding HIV), and employment as an essential worker including both healthcare and non-healthcare essential workers (defined as those working in retail, transportation, social services, and other services deemed essential), (36) all assessed by self-report.

Among PLWH, we measured median time living with HIV (median [IQR]), the proportion on ART, with an undetectable HIV viral load (<50 copies/mL), receipt of HIV medical care since the COVID-19 restrictions were implemented in mid-March 2020, and how much their HIV status affected their fear of acquiring COVID-19 (more/much more fearful *vs* no difference *vs* less/much less fearful).

### WHO Vaccine hesitancy scales and psychological constructs within the Theory of Planned Behavior

The questionnaire assessed several psychological constructs as potential correlates of vaccine intention, including (1) a modified ***WHO Vaccine Hesitancy Scale*** (16,25), which included two factors: ***Lack of Vaccine Confidence*** (7-item 5-point Likert scale from Strongly Agree to Strongly Disagree, with higher agreement corresponding with higher lack of general vaccine confidence) and ***Vaccine Risk*** (2-item 5-point Likert scale from Strongly Agree to Strongly Disagree, with higher agreement corresponding with higher concerns about vaccine risks); and grounded in the Theory of Planned Behavior (26), items developed and previously used to measure key factors shown to influence COVID-19 vaccine intention (19) including (2) ***Attitudes towards the COVID-19 vaccine*** (8 item 5-point Likert scale from Strongly Agree to Strongly Disagree, with higher agreement corresponding with more positive attitudes towards the COVID-19 vaccine); (3) **Perceived Behavioral Control** to receive a COVID-19 vaccine (4 item 5-point Likert scale from Strongly Agree to Strongly Disagree, with higher agreement corresponding with higher self-perception of being able to receive the COVID-19 vaccine if desired); (4) the influence of ***Direct Social Norms*** (4-item 5-point Likert scale from Strongly Agree to Strongly Disagree, with higher agreement corresponding with being more likely to be influenced by direct social norms to receive the COVID-19 vaccine); and (5) the influence of ***Indirect Social Norms*** (8 item 5-point Likert scale assessing both whether various influencers would Strongly Approve to Strongly Disapprove of the participant receiving the COVID-19 vaccine and how much the participant Strongly Agrees to Strongly Disagrees that what the influencer thinks is important to them, with higher scores indicating a greater influence of indirect social norms). All scale items are shown in **Table IV**.

### Data Analyses

Descriptive statistics (mean (Standard Deviation (±SD)) or median [IQR] for continuous variables and n (%) for categorical variables) were used to characterize baseline distributions of study variables, stratified by HIV status. Baseline differences were compared using Wilcoxon rank sum test for continuous variables and Fisher’s exact test for categorical variables.

Descriptive statistics were also used to report the prevalence of intention to vaccinate by HIV status. Bivariable analyses examined the relationship between intention to vaccinate and socio-demographic variables. An exploratory multivariable logistic regression model was used to examine the crude and adjusted odds ratios (with 95% confidence intervals) between HIV status and vaccine intention controlling for potential socio-demographic confounders. After assessing collinearity, *a priori* possible predictors of vaccine intention with P<0.1 in bivariable analyses were considered in the multivariable model. Multivariable analyses included only non-missing data.

For each of the items in the WHO Vaccine Hesitancy Scale and the psychological constructs, the proportion of participants reporting Strongly Agree/Agree (vs Neutral/Disagree/Strongly Disagree) were reported and compared by HIV status, with differences compared using Pearson χ2 or Fisher’s exact test. We also computed the mean (±SD) total score of each scale and compared means by HIV status using Wilcoxon rank sum tests.

Among PLWH, we used logistic regression to examine associations between socio-demographic characteristics, the WHO Vaccine Hesitancy Scale, and the psychological constructs with COVID-19 vaccine intention.

All p-values were two-sided and considered statistically significant at P<0.05. Analyses were conducted in R v.4.0.2 (37).

## RESULTS

Between August 20^th^, 2020 and March 1^st^, 2021, 6,518 individuals completed the online survey, of whom 5,588 (85.7%) identified as women or gender diverse individuals and were included in this analysis. Of these, 69 (1.23%) were living with HIV (LWH) whereas 5,519 (98.8%) were not LWH.

### Baseline characteristics

Age was similar among participants LWH (mean ± SD: 49.9 ± 11.4 years) and not LWH (48.1 ± 12.1 years) and a majority reported being assigned female sex at birth (99.6%). Participants LWH reported significantly greater gender, ethno-racial, and socio-economic diversity. Compared to those not LWH, individuals LWH were significantly more likely to identify as gender diverse (8.7% vs 1.2%; p=0.0003), of Indigenous ancestry (29% vs 3%; p<0.001), African Caribbean or Black (8.7% vs 0.3%; p<0.0001), report a household income below $20,000/year (17.4% vs 2.3%; p<0.0001), and a highschool education or less (34.8% vs 12.2%; p<0.0001). There were no differences by essential worker employment (27.5% vs 32.9%; p=0.30) (**Table I**).

**Table I:**
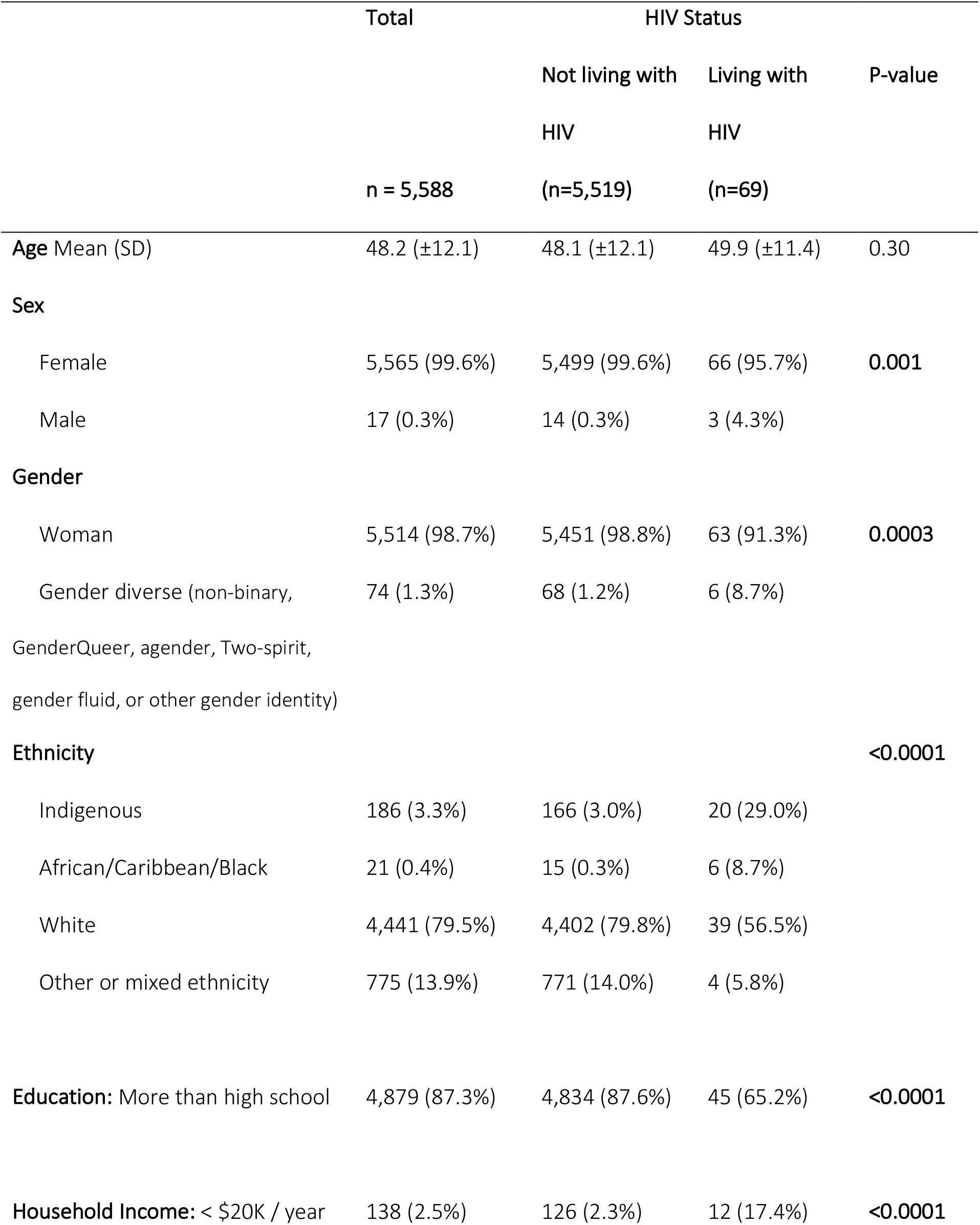
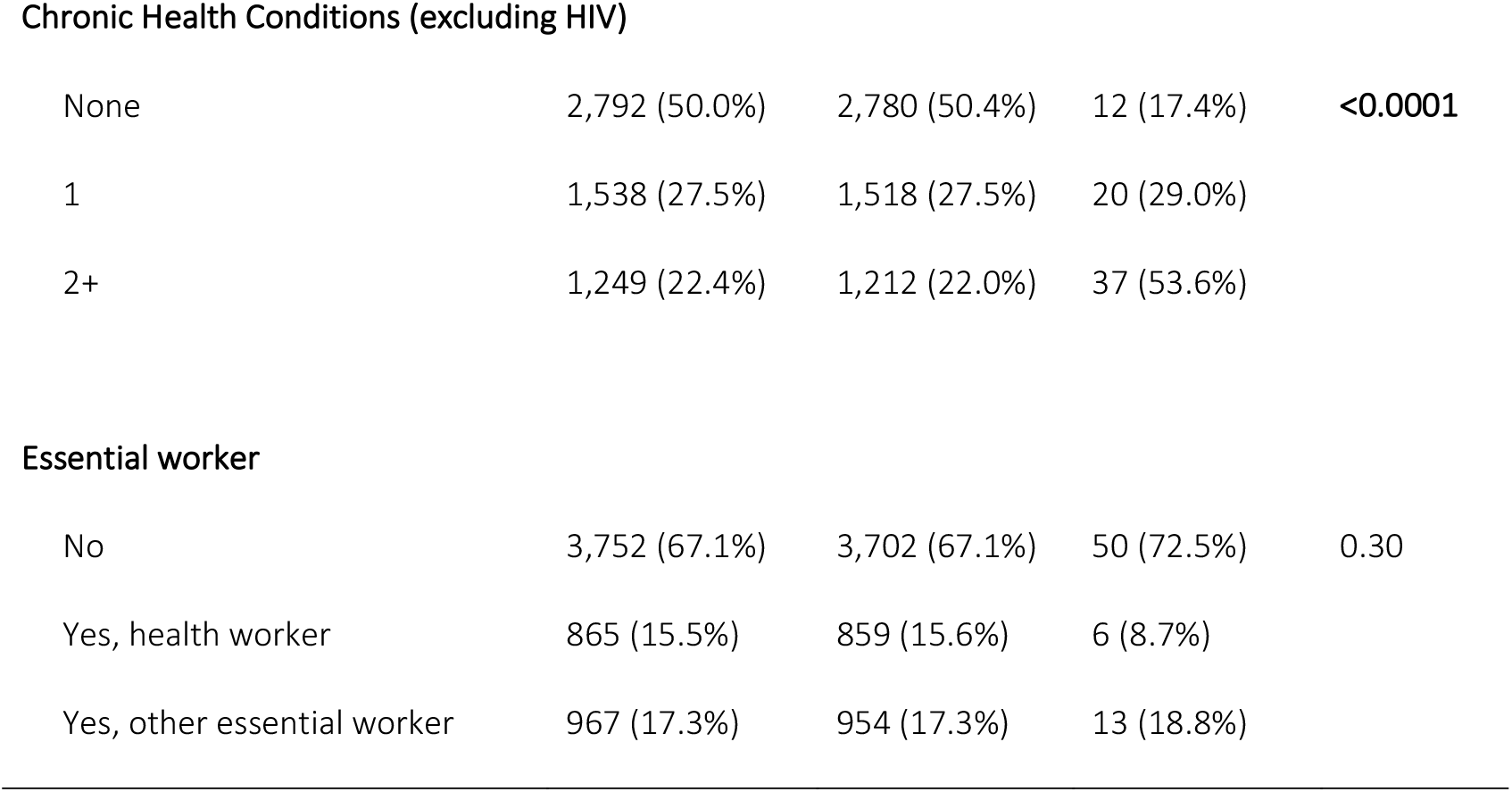
Baseline Characteristics of study sample overall and by HIV status, column % (n=5,588)

|  | Total | HIV Status |  |  |
| --- | --- | --- | --- | --- |
|  |  | Not living with<br>HIV | Living with<br>HIV | P-value |
|  | n = 5,588 | (n=5,519) | (n=69) |  |
| **Age** Mean (SD) | 48.2 (±12.1) | 48.1 (±12.1) | 49.9 (±11.4) | 0.30 |
| **Sex** |  |  |  |  |
| Female | 5,565 (99.6%) | 5,499 (99.6%) | 66 (95.7%) | **0.001** |
| Male | 17 (0.3%) | 14 (0.3%) | 3 (4.3%) |  |
| **Gender** |  |  |  |  |
| Woman | 5,514 (98.7%) | 5,451 (98.8%) | 63 (91.3%) | **0.0003** |
| Gender diverse (non-binary,<br>GenderQueer, agender, Two-spirit,<br>gender fluid, or other gender identity) | 74 (1.3%) | 68 (1.2%) | 6 (8.7%) |  |
| **Ethnicity** |  |  |  | **<0.0001** |
| Indigenous | 186 (3.3%) | 166 (3.0%) | 20 (29.0%) |  |
| African/Caribbean/Black | 21 (0.4%) | 15 (0.3%) | 6 (8.7%) |  |
| White | 4,441 (79.5%) | 4,402 (79.8%) | 39 (56.5%) |  |
| Other or mixed ethnicity | 775 (13.9%) | 771 (14.0%) | 4 (5.8%) |  |
| **Education:** More than high school | 4,879 (87.3%) | 4,834 (87.6%) | 45 (65.2%) | **<0.0001** |
| **Household Income:** < \$20K / year | 138 (2.5%) | 126 (2.3%) | 12 (17.4%) | **<0.0001** |
| **Chronic Health Conditions (excluding HIV)** |  |  |  |  |
| None | 2,792 (50.0%) | 2,780 (50.4%) | 12 (17.4%) | **<0.0001** |
| 1 | 1,538 (27.5%) | 1,518 (27.5%) | 20 (29.0%) |  |
| 2+ | 1,249 (22.4%) | 1,212 (22.0%) | 37 (53.6%) |  |
| **Essential worker** |  |  |  |  |
| No | 3,752 (67.1%) | 3,702 (67.1%) | 50 (72.5%) | 0.30 |
| Yes, health worker | 865 (15.5%) | 859 (15.6%) | 6 (8.7%) |  |
| Yes, other essential worker | 967 (17.3%) | 954 (17.3%) | 13 (18.8%) |  |

A higher proportion of participants LWH reported living with ≥1 chronic health condition (excluding HIV) (82.6% vs 49.6%; p<0.0001) and were significantly more likely to report living with chronic obstructive pulmonary disease (COPD) or emphysema, chronic lung disease, heart disease, liver disease and liver cirrhosis, and renal problems compared with those not LWH.

### Characteristics of participants living with HIV

Median years living with HIV was 20.5 [IQR: 14-17], 79.7% were currently on ART for a median of 14.0 years [10-23 years], and 73.9% reported being virally undetectable (<50 copies/mL). Overall, 62.3% reported receiving any HIV medical care since the COVID-19 restrictions were implemented and 58.6% reported that their HIV-positive status made them more fearful of acquiring COVID-19 (3.4% reported less fearful, 37.9% reported that it makes no difference) (**Table II**).

**Table II:** Baseline characteristics of people living with HIV enrolled in the RESPPONSE study (n=69)

| Characteristic | n or Median | % or IQR |
| --- | --- | --- |
| Median years living with HIV | 20.5 | 14.0-17.0 |
| Currently on antiretroviral therapy (ART) | 55 | 79.7% |
| Median years on ART | 14.0 | 10.0-23.0 |
| Undetectable HIV viral load (<50 copies/mL) | 51 | 73.9% |
| Received any HIV medical care since COVID-19 restrictions | 43 | 62.3% |
| How much does your HIV status affect your fear of acquiring COVID-19? (n=58) |  |  |
| More/much more fearful | 34 | 58.6% |
| It makes no difference | 22 | 37.9% |
| Less/much less fearful | 2 | 3.4% |

### Intention to receive a COVID-19 vaccine by HIV status and socio-demographic characteristics

In the overall sample, 79.7% reported being “very or somewhat likely” to receive a COVID-19 vaccine if it were to become available to the public and recommended for them. Intention to vaccinate was significantly lower among participants LWH compared with those not LWH (65.2% vs 79.6%; p=0.009. OR: 0.49; 95%CI:0.30-0.83). (**Table III**)

**Table III:**
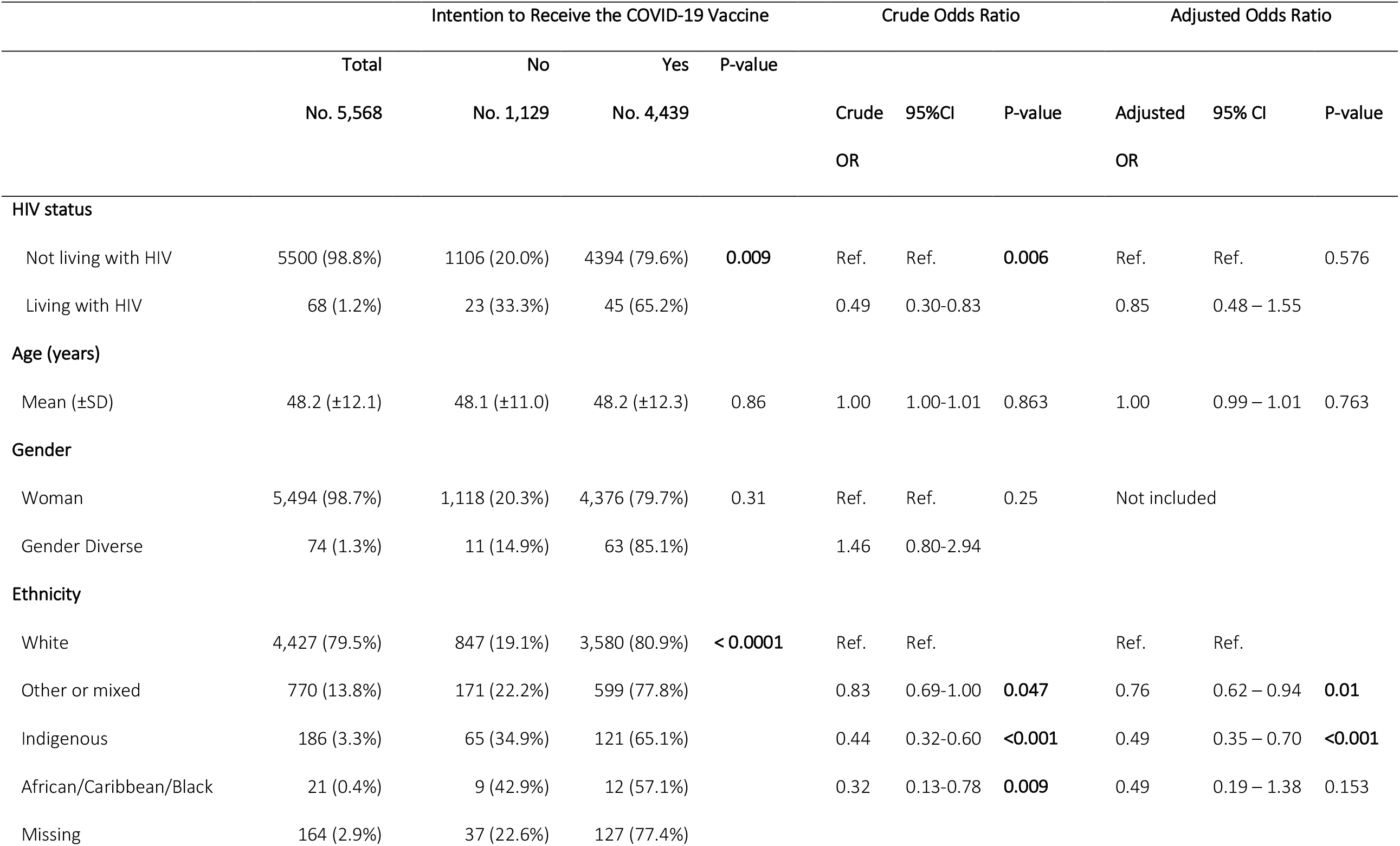
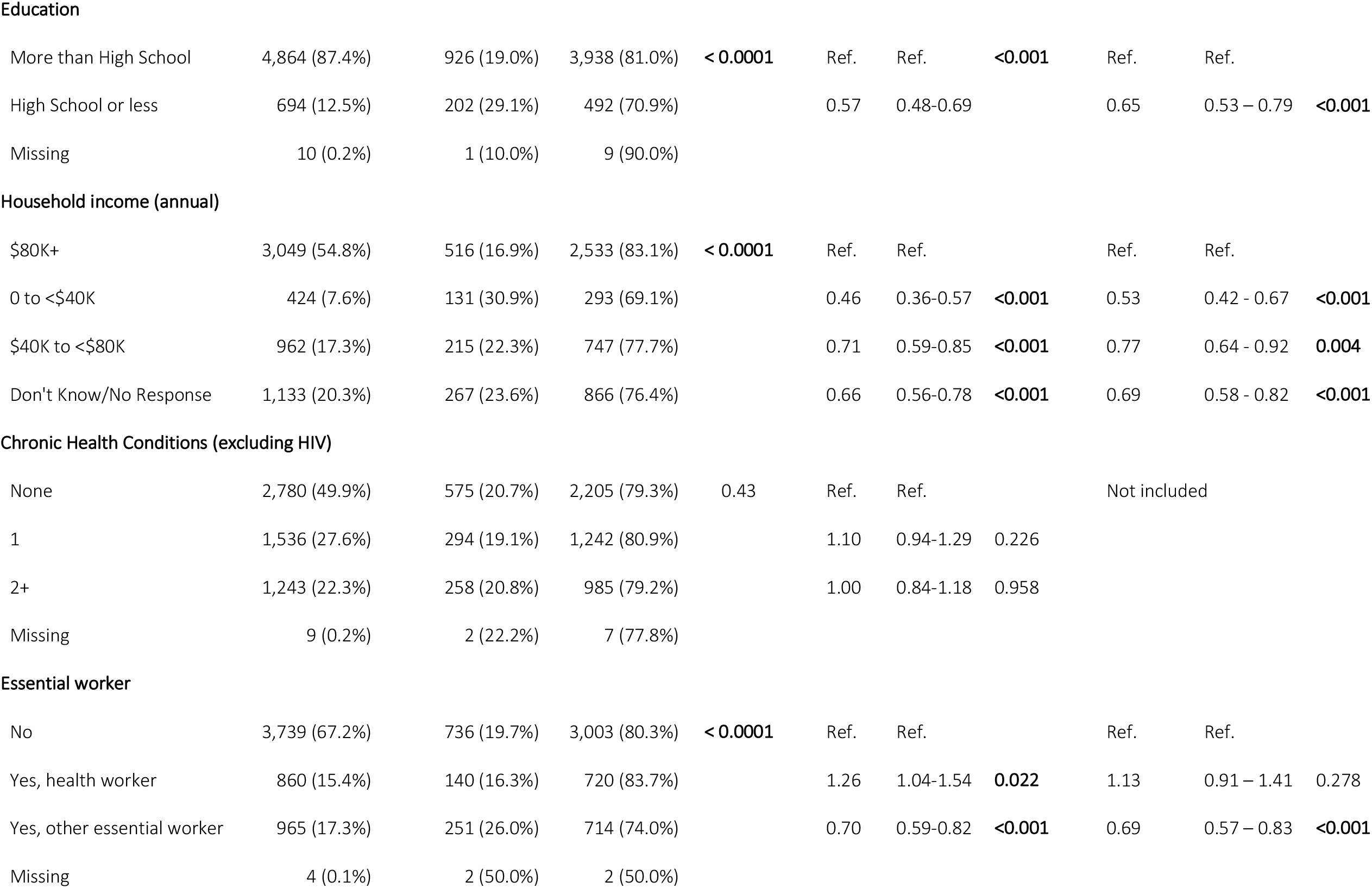
Unadjusted and Adjusted Odds Ratios (and 95% Confidence Intervals) of Vaccine Intention by HIV status and socio-demographic variables, row %.

|  | Intention to Receive the COVID-19 Vaccine |  |  |  | Crude Odds Ratio |  |  | Adjusted Odds Ratio |  |  |
| --- | --- | --- | --- | --- | --- | --- | --- | --- | --- | --- |
|  | Total | No | Yes | P-value |  |  |  |  |  |  |
|  | No. 5,568 | No. 1,129 | No. 4,439 |  | Crude | 95%CI | P-value | Adjusted | 95% CI | P-value |
|  |  |  |  |  | OR |  |  | OR |  |  |
| HIV status |  |  |  |  |  |  |  |  |  |  |
| Not living with HIV | 5500 (98.8%) | 1106 (20.0%) | 4394 (79.6%) | 0.009 | Ref. | Ref. | 0.006 | Ref. | Ref. | 0.576 |
| Living with HIV | 68 (1.2%) | 23 (33.3%) | 45 (65.2%) |  | 0.49 | 0.30-0.83 |  | 0.85 | 0.48 – 1.55 |  |
| Age (years) |  |  |  |  |  |  |  |  |  |  |
| Mean (±SD) | 48.2 (±12.1) | 48.1 (±11.0) | 48.2 (±12.3) | 0.86 | 1.00 | 1.00-1.01 | 0.863 | 1.00 | 0.99 – 1.01 | 0.763 |
| Gender |  |  |  |  |  |  |  |  |  |  |
| Woman | 5,494 (98.7%) | 1,118 (20.3%) | 4,376 (79.7%) | 0.31 | Ref. | Ref. | 0.25 | Not included |  |  |
| Gender Diverse | 74 (1.3%) | 11 (14.9%) | 63 (85.1%) |  | 1.46 | 0.80-2.94 |  |  |  |  |
| Ethnicity |  |  |  |  |  |  |  |  |  |  |
| White | 4,427 (79.5%) | 847 (19.1%) | 3,580 (80.9%) | < 0.0001 | Ref. | Ref. |  | Ref. | Ref. |  |
| Other or mixed | 770 (13.8%) | 171 (22.2%) | 599 (77.8%) |  | 0.83 | 0.69-1.00 | 0.047 | 0.76 | 0.62 – 0.94 | 0.01 |
| Indigenous | 186 (3.3%) | 65 (34.9%) | 121 (65.1%) |  | 0.44 | 0.32-0.60 | <0.001 | 0.49 | 0.35 – 0.70 | <0.001 |
| African/Caribbean/Black | 21 (0.4%) | 9 (42.9%) | 12 (57.1%) |  | 0.32 | 0.13-0.78 | 0.009 | 0.49 | 0.19 – 1.38 | 0.153 |
| Missing | 164 (2.9%) | 37 (22.6%) | 127 (77.4%) |  |  |  |  |  |  |  |
| Education |  |  |  |  |  |  |  |  |  |  |
| More than High School | 4,864 (87.4%) | 926 (19.0%) | 3,938 (81.0%) | **< 0.0001** | Ref. | Ref. | **<0.001** | Ref. | Ref. |  |
| High School or less | 694 (12.5%) | 202 (29.1%) | 492 (70.9%) |  | 0.57 | 0.48-0.69 |  | 0.65 | 0.53 – 0.79 | **<0.001** |
| Missing | 10 (0.2%) | 1 (10.0%) | 9 (90.0%) |  |  |  |  |  |  |  |
| Household income (annual) |  |  |  |  |  |  |  |  |  |  |
| \$80K+ | 3,049 (54.8%) | 516 (16.9%) | 2,533 (83.1%) | **< 0.0001** | Ref. | Ref. | | Ref. | Ref. | |
| 0 to <\$40K | 424 (7.6%) | 131 (30.9%) | 293 (69.1%) | | 0.46 | 0.36-0.57 | **<0.001** | 0.53 | 0.42 - 0.67 | **<0.001** |
| \$40K to <\$80K | 962 (17.3%) | 215 (22.3%) | 747 (77.7%) | | 0.71 | 0.59-0.85 | **<0.001** | 0.77 | 0.64 - 0.92 | **0.004** |
| Don't Know/No Response | 1,133 (20.3%) | 267 (23.6%) | 866 (76.4%) |  | 0.66 | 0.56-0.78 | **<0.001** | 0.69 | 0.58 - 0.82 | **<0.001** |

Chronic Health Conditions (excluding HIV)
|  |  |  |  |  |  |  |  |  |
| --- | --- | --- | --- | --- | --- | --- | --- | --- |
| None | 2,780 (49.9%) | 575 (20.7%) | 2,205 (79.3%) | 0.43 | Ref. | Ref. |  | Not included |
| 1 | 1,536 (27.6%) | 294 (19.1%) | 1,242 (80.9%) |  | 1.10 | 0.94-1.29 | 0.226 |  |
| 2+ | 1,243 (22.3%) | 258 (20.8%) | 985 (79.2%) |  | 1.00 | 0.84-1.18 | 0.958 |  |
| Missing | 9 (0.2%) | 2 (22.2%) | 7 (77.8%) |  |  |  |  |  |
| Essential worker |  |  |  |  |  |  |  |  |  |  |
| No | 3,739 (67.2%) | 736 (19.7%) | 3,003 (80.3%) | **< 0.0001** | Ref. | Ref. |  | Ref. | Ref. |  |
| Yes, health worker | 860 (15.4%) | 140 (16.3%) | 720 (83.7%) |  | 1.26 | 1.04-1.54 | **0.022** | 1.13 | 0.91 – 1.41 | 0.278 |
| Yes, other essential worker | 965 (17.3%) | 251 (26.0%) | 714 (74.0%) |  | 0.70 | 0.59-0.82 | **<0.001** | 0.69 | 0.57 – 0.83 | **<0.001** |
| Missing | 4 (0.1%) | 2 (50.0%) | 2 (50.0%) |  |  |  |  |  |  |  |

**Table IV.**
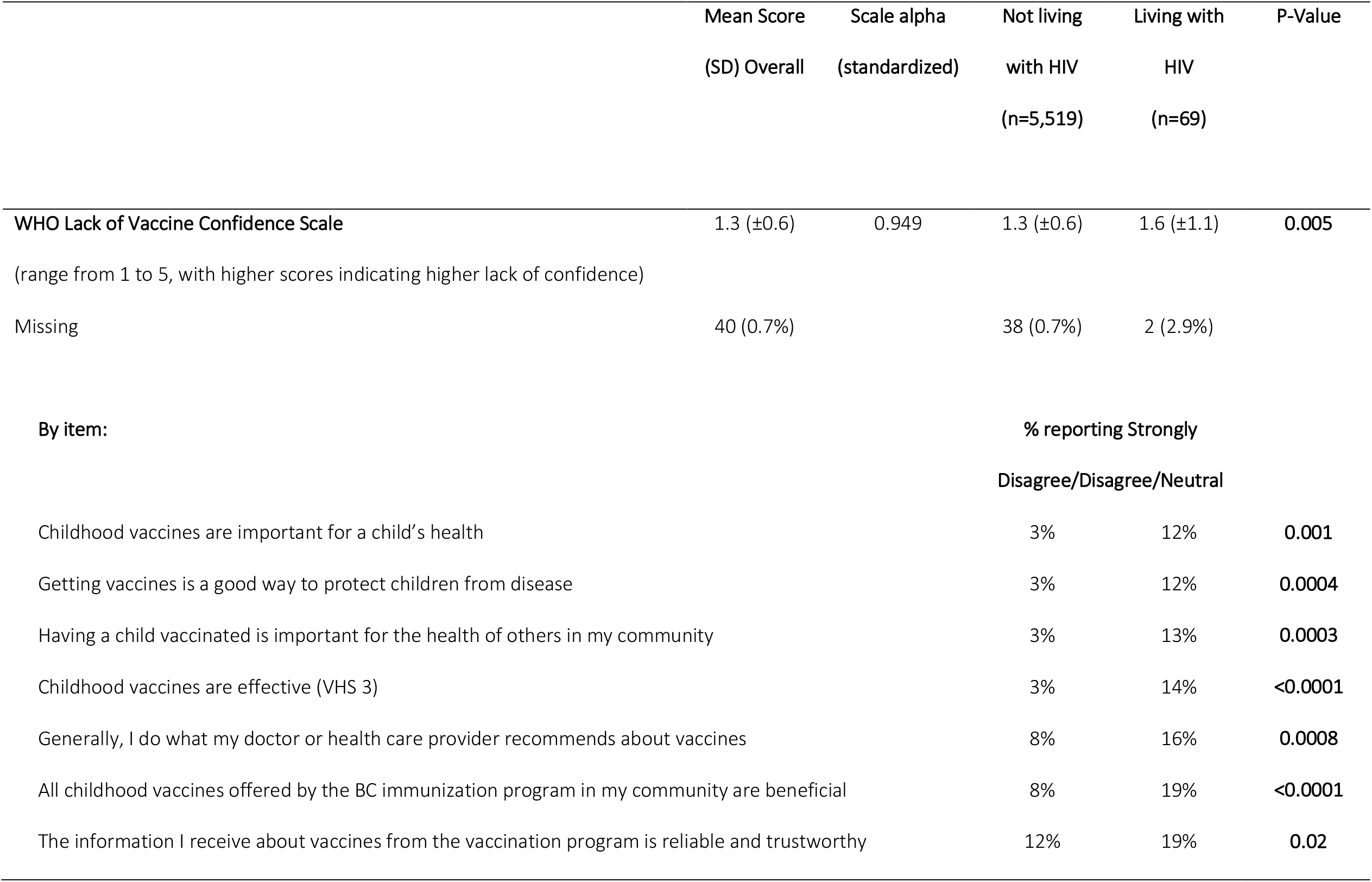
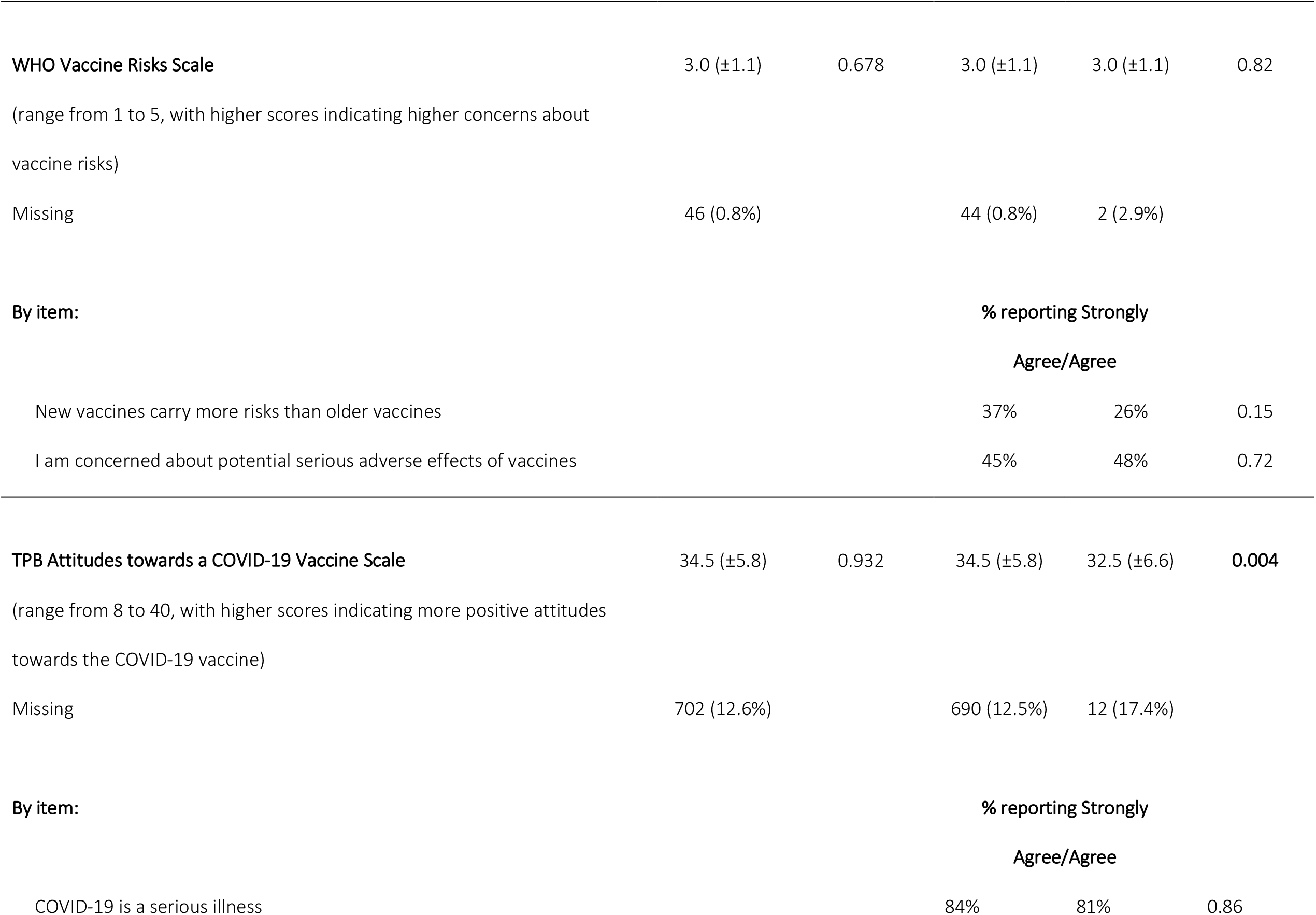
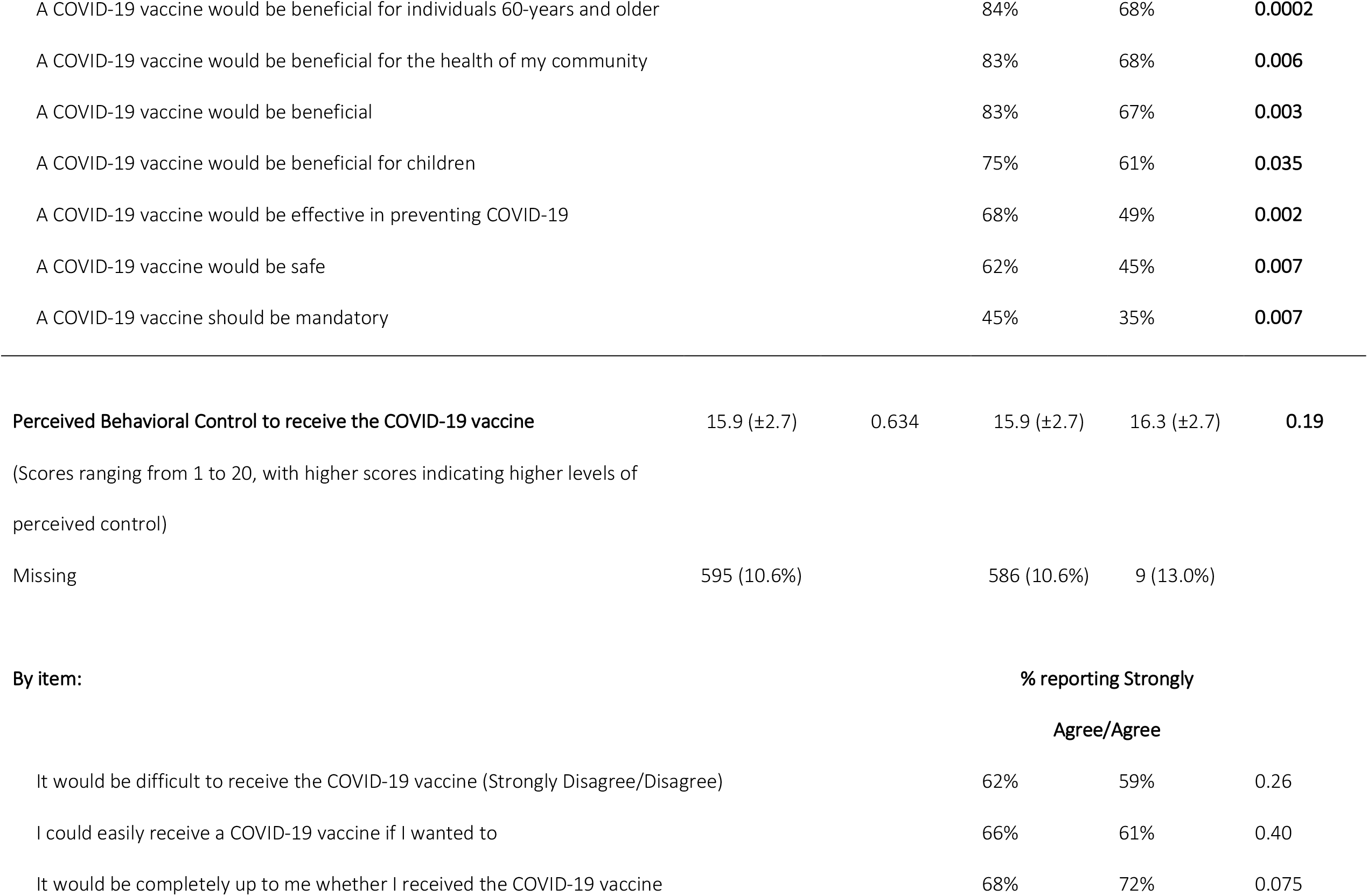
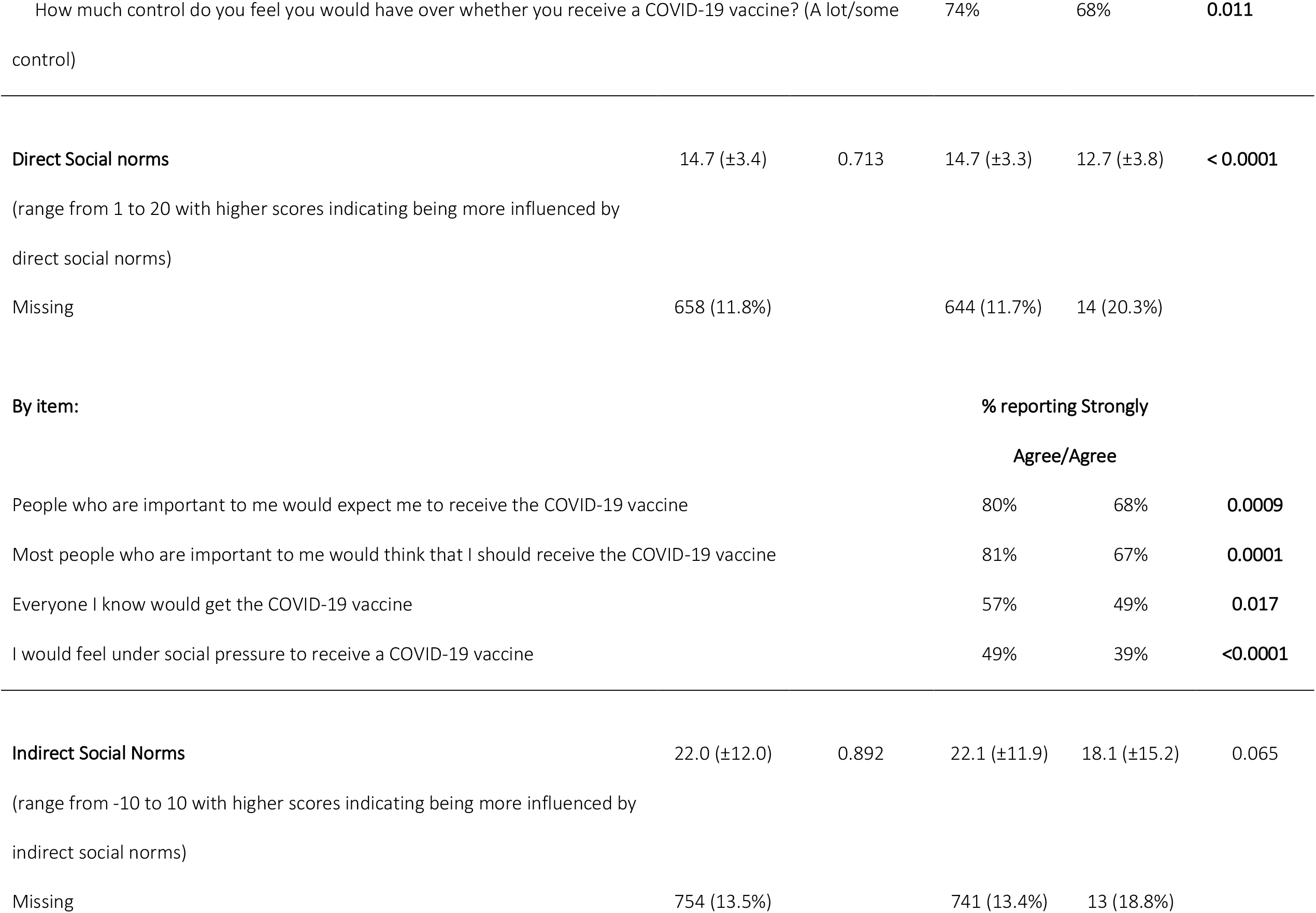
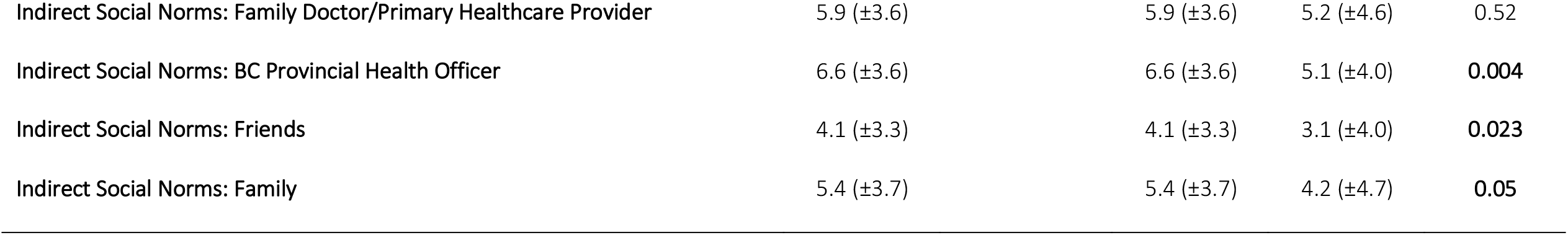
Vaccine Hesitancy and COVID-19 vaccine Psychological Constructs by HIV status, column %.

In the full sample, intention to vaccinate was also lower among racialized individuals, including people of Indigenous ancestry (65.1%; OR: 0.44; 95% CI: 0.32-0.60), African/Caribbean/and Black (57.1%; OR: 0.32; 95% CI: 0.13-0.78), and people of other or mixed ethnicities (77.8%; OR: 0.83; 95% CI: 0.69-1.00) relative to white participants. Participants residing in lower income households, with less education, or essential workers not in the health sector were also significantly less likely to report an intention to vaccinate. There were no significant differences by age, gender, or the presence of chronic health conditions.

In the multivariable model, living with HIV was no longer significantly associated with intention to vaccinate (adjusted OR:0.85; 95%CI: 0.48-1.55). The observed effect in unadjusted analyses was attenuated by differences in the distribution of ethnicity, household income, education, and essential worker status between groups. Compared to white participants, people of Indigenous ancestry (aOR: 0.49; 95% CI: 0.35-0.70) and people of other or mixed ethnicities (aOR: 0.76; 95% CI: 0.62-0.94) had significantly lower adjusted odds of reporting an intention to vaccinate. There was no significant difference among African/Caribbean/and Black participants, although the sample was small (OR: 0.49; 95% CI: 0.19-1.38). Participants residing in lower income households (<$40K per year aOR: 0.53; 95% CI: 0.42-0.67 and $40K to <$80K per year aOR: 0.77; 95% CI: 0.64-0.92 compared with those with household incomes of ≥$80K per year), with a high school education or less (aOR: 0.65; 95% CI: 0.53-0.79), or who were essential workers not in the health sector (aOR: 0.69; 95% CI: 0.57-0.83) had significantly lower adjusted odds of reporting an intention to vaccinate.

### WHO Vaccine Hesitancy Scale and Psychological Constructs by HIV status

All scales demonstrated good to strong agreement (Cronbach’s alpha ranging from a low of 0.63 for the Perceived Behavioral Control scale to a high of 0.95 for the WHO Lack of Vaccine Confidence Scale) (**Table V**).

**Table V.** Bivariable associations between socio-demographic, vaccine hesitancy, and psychological constructs and intention to receive the COVID-19 vaccine among women and gender diverse individuals living with HIV (n=69)

|  | Crude OR | 95% CI | P-value |
| --- | --- | --- | --- |
| **Age (per year increase)** | 1.05 | 1.00-1.10 | **0.048** |
| **Indigenous Ancestry** |  |  |  |
| No | Ref. | Ref. |  |
| Yes | 0.38 | 0.12-1.19 | 0.10 |
| **Racialized** |  |  |  |
| No (White) | Ref. | Ref. |  |
| Yes (Indigenous, African/Caribbean/Black, Other/mixed ethnicity) | 0.63 | 0.20-2.03 | 0.43 |
| **Education** |  |  |  |
| More than High School | Ref. | Ref. |  |
| High School or less | 0.40 | 0.14-1.13 | 0.09 |
| **Household Income** |  |  |  |
| \$20K+ per year | Ref. | Ref. | |
| <\$20K per year | 0.43 | 0.11-1.62 | 0.21 |
| Don't Know/No Answer | 0.71 | 0.15-3.93 | 0.68 |
| **Chronic Health Conditions** |  |  |  |
| None | Ref. | Ref. |  |
| 1 or more | 3.50 | 0.98-13.4 | **0.05** |
| **Essential worker** |  |  |  |
| No | Ref. | Ref. |  |
| Yes, health worker | 0.44 | 0.07-2.62 | 0.35 |
| Yes, other essential worker | 0.71 | 0.20-2.66 | 0.59 |
| WHO Lack of Vaccine Confidence Scale | 0.40 | 0.18 – 0.71 | **0.007** |
| WHO Vaccine Risks Scale | 0.69 | 0.41 – 1.11 | 0.141 |
| Attitudes toward the COVID-19 Vaccine Scale | 1.31 | 1.15 – 1.54 | **<0.001** |
| Perceived Behavioral Control Scale | 1.23 | 1.00 – 1.53 | 0.058 |
| Direct Social Norms Scale | 1.27 | 1.08 – 1.54 | **0.007** |
| Indirect Social Norms: Total Scale | 1.16 | 1.09 – 1.28 | **<0.001** |
| Indirect Social Norms: Family Doctor/Primary Healthcare Provider | 1.31 | 1.13 – 1.55 | **0.001** |
| Indirect Social Norms: BC Provincial Health Officer | 1.74 | 1.36 – 2.48 | **<0.001** |
| Indirect Social Norms: Friends | 1.58 | 1.25 – 2.20 | **0.001** |
| Indirect Social Norms: Family | 1.65 | 1.32 – 2.25 | **<0.001** |

*Lack of vaccine confidence* was low overall, however, participants LWH expressed significantly higher lack of vaccine confidence (or higher vaccine hesitancy) across each of the 7 scale items. Among those LWH, mean Lack of Vaccine Confidence score was 1.6 (SD=1.1) compared with 1.3 (SD=0.6) among those not LWH (p=0.005).

*Perceptions of vaccine risks* were higher overall, however, there were no significant differences by HIV status. Among respondents LWH, the mean Vaccine Risk score was 3.0 (SD=1.1) and highly similar to that among those not LWH (3.0 (SD=1.1), p=0.82).

*Attitudes towards the COVID-19 vaccine* were positive with at least 75% Strongly Agreeing/ Agreeing with most scale items, with the exception of three items where a lower proportion of participants agreed that a COVID-19 vaccine would be effective at preventing COVID-19, would be safe, or should be mandatory. Participants LWH expressed significantly less positive attitudes towards the COVID-19 vaccine across each of the 8 scale items, with the exception of “COVID-19 is a serious illness” where agreement was similar (81.1% vs 84.2%; p=0.86). Among participants LWH, the mean Attitudes towards the COVID-19 vaccine score was 32.5 (SD=6.6) compared with 34.5 (SD=5.8) among those not LWH (p=0.004).

Approximately two-thirds of participants reported *perceiving that they had high behavioral control* over whether or not they could receive the COVID-19 vaccine if they wanted to, with no overall differences in the scale score by HIV status (p=0.19).

Overall, participants were influenced by *direct social norms* to receive a COVID-19 vaccine. A large majority agreed that people who are important to them would expect them to receive the COVID-19 vaccine and think that they should receive the COVID-19 vaccine. Agreement for the other two scale items was lower, including “Everyone I know would get the COVID-19 vaccine” and feeling “under social pressure to receive the COVID-19 vaccine”. Participants LWH were significantly less likely to be influenced by direct social norms to receive the COVID-19 vaccine than those not LWH (mean score: 12.7 *vs* 14.7, respectively; p<0.0001).

Participants were similarly likely to report being influenced by *indirect social norms* overall, however, participants LWH were significantly less likely to be influenced by the BC Provincial Health Officer (the senior public health official directing the COVID-19 public health response), friends, or family. They were equally likely as participants not LWH to report being influenced by their family doctor/primary healthcare provider (PHCP). Among participants LWH, the mean total Indirect Social Norms score was 18.1 (SD=15.2) compared with 22.1 (SD=11.9) among those not LWH (p=0.065).

### Predictors of Intention to Vaccinate among participants living with HIV

All the psychological constructs were significantly associated with vaccine intention in the overall sample, as expected and as previously shown (19) (all p-values <0.001). **(Supplementary Table I)**.

Participants LWH who had a higher odds of reporting an intention to vaccinate were older (OR: 1.05 per year increase; 95% CI: 1.00-1.10), reported one or more chronic health conditions (OR: 3.50; 95% CI: 0.98-13.43), had lower lack of vaccine confidence (0.40; 95%CI: 0.18-0.71) more positive attitudes towards the COVID-19 vaccine (OR: 1.31; 95% CI: 1.15-1.54), greater influence of direct social norms (OR: 1.27; 95% CI: 1.08-1.54), and greater influence of indirect social norms from family doctors/PHCPs (OR: 1.31; 95% CI: 1.13-1.55), the BC provincial health officer (OR: 1.74; 95CI%: 1.36-2.48), friends (OR: 1.58; 95%CI: 1.25-2.20), and family (OR: 1.65; 95%CI: 1.32-2.25). There was no statistically significant association between intention to vaccinate and perceived vaccine risks, perceived behavioral control, other assessed socio-demographic variables (ethnicity, education, household income, essential worker status), or perceived risk of acquiring COVID-19 due to HIV status. Owing to missing data and small cell sizes, we were not able to assess associations with HIV clinical variables (ART use, undetectable viral load).

Vaccine confidence demonstrated the largest effect, whereby participants LWH who expressed vaccine confidence had 2.5 fold higher odds of vaccine intention compared with those who lacked vaccine confidence. Given the small sample size and high degree of correlation between psychological constructs, adjusted analyses were not performed.

## DISCUSSION

In this large population-based sample of women and gender diverse individuals in BC, we found that only two-thirds (65.2%) of participants LWH reported intending to receive a COVID-19 vaccine if recommended and available to them, significantly lower than participants not LWH (79.6%). HIV status itself, however, was not significantly associated with COVID-19 vaccine intention in adjusted analyses. This finding is illustrative of the wide gap between the strong recommendations for COVID-19 vaccination for all PLWH and current intentions (4,10,14). Findings are further concerning given the large proportion of participants LWH who belong to other communities prioritized for vaccine receipt due to higher risk of COVID-19 infection and severe illness, including those experiencing social inequities and co-morbidities.

The observed effect of HIV status on vaccine intention in unadjusted analyses was explained by differences in the distribution of other key socio-demographic factors, including Indigenous ancestry, being racialized, lower household income, lower education, and essential worker (non-health related) status, all previously shown to be associated with vaccine intention in the general BC population (19). These findings are consistent with research from two general population studies in the US which reported nearly 80% of participants overall were likely/somewhat likely to receive the COVID-19 vaccine, with significantly lower prevalence among racialized and lower socio-economic status participants (18,38).

We also found significant differences in vaccine hesitancy and psychological constructs that shape vaccine intention and uptake behaviors by HIV status. Participants LWH reported lower vaccine confidence, less positive attitudes towards the COVID-19 vaccine, and were less likely to be influenced by direct or indirect social norms to receive the COVID-19 vaccine. These findings are consistent with findings from a US study of Black Americans living with HIV who reported widespread COVID-19 mistrust, with over half reporting COVID-19 vaccine hesitancy (17).

Findings further suggest that efforts to address vaccine confidence and the psychological constructs measured using the Theory of Planned Behavior are important for supporting vaccine intention and uptake. Among participants LWH, we found that differences in the social determinants of health did not predict vaccine intentions. Rather, those with higher vaccine confidence, positive attitudes toward the COVID-19 vaccine, and those who were more strongly influenced by direct and indirect social norms had significantly higher odds of reporting vaccine intention. Collectively, these data suggest that targeted and consistent messaging from family doctors/PHCPs and senior public health officials stating that COVID-19 vaccines are safe, effective, beneficial, and strongly recommended for PLWH, may be a pathway to improve vaccine confidence and attitudes. Specific information relevant for PLWH includes data regarding the immunogenicity and safety of COVID-19 vaccines among PLWH (39) and the importance of maintaining engagement in HIV care and adhering to ART even among those who are vaccinated. For reproductive-aged WLWH seeking to conceive, evidence regarding the safety and effectiveness of COVID-19 vaccines during pregnancy and breastfeeding will be further reassuring (40). The opportunity for HIV care provider-led discussions is particularly relevant given that a majority of PLWH are engaged in HIV medical care and over half expressed being fearful that their HIV status would affect their risk of acquiring COVID-19. Research has shown that WLWH express high trust in their HIV care providers and identify them as the preferred source of relevant non-HIV specific information (41). As we collectively move into the next “ground game” phase of increasing vaccine coverage, public health campaigns will need to support and foster these trusting relationships.

Efforts to support vaccine decision-making and uptake among PLWH can benefit from adopting community-based research principles of meaningful community involvement and engagement across the COVID-19 vaccine response (42,43). Research with WLWH, has highlighted the profound influence that peers have on increasing knowledge and healthcare support for WLWH and these learnings should be extended to support informed vaccine decision-making and uptake of a COVID-19 vaccine.

The prevalence of vaccine intention over the course of data collection (August 2020-March 1, 2021) corresponds with a period of time during which the COVID-19 vaccine was not widely available in Canada (10). Beginning in April 2021, vaccine eligibility expanded from priority groups to the general population, beginning with older individuals and extending to those aged 12+ years by June 2021 (44). As of July 26, 2021, 81% of BC residents aged 12+ years were partially vaccinated while 62% were fully vaccinated (45). This first-dose vaccine uptake rate is highly consistent with our estimate of 79.7% of adults reporting vaccine intention. While no provincial estimates are yet available for PLWH, these data provide external validity to our findings.

### Limitations

Although the number of participants LWH was small, the proportion of those LWH in this sample was higher than expected through general population-based recruitment strategies (46), enabling comparisons with the general population. We did not have sufficient sample size to conduct separate analyses for women and gender diverse individuals LWH. However, in bivariable analyses we did not observe significant differences in vaccine intention by gender, which further informed our decision to include both women and gender diverse individuals in analyses. The study sample was drawn from individuals who were sufficiently concerned about COVID-19, reasonably trusting of scientific research, and with sufficient resources (technological, time) to complete the online survey. Thus, our findings may over-estimate the true prevalence of vaccine intention.

## CONCLUSION

Among a sample of women and gender diverse individuals, we found important disparities in vaccine intentions by HIV status. Vaccine intentions are, however, dynamic and may evolve as vaccine delivery programs expand. Ongoing efforts must ensure that people living with HIV, and other historically marginalized populations, continue to have equitable access to vaccines and up-to-date vaccine information. Such efforts must acknowledge that the same socio-structural inequities and injustices that produce HIV risk and consequence for women and gender diverse people undermine vaccine confidence and fuel COVID-19 inequities. Our findings suggest pathways for building vaccine confidence, address vaccine concerns, and support informed vaccination decision-making. In partnership with communities, such pathways can be leveraged to promote COVID-19 vaccine uptake among women and gender diverse people living with HIV.

## Data Availability

Data used in this analysis will be available on the Womens Health Research Institute (WHRI) website.

https://www.whri.org

## REFERENCES

1. Government of Canada. COVID-19 daily epidemiology update. 2021; https://health-infobase.canada.ca/covid-19/epidemiological-summary-covid-19-cases.html#a7. Accessed July 10, 2021. Accessed Accessed July 10, 2021, Accessed July 10, 2021.

2. Sundaram ME, Calzavara A, Mishra S, et al. Individual and social determinants of SARS-CoV-2 testing and positivity in Ontario, Canada: a population-wide study. CMAJ. May 17 2021;193(20):E723–E734.

3. McKenzie K. Socio-demographic data collection and equity in covid-19 in Toronto. EClinicalMedicine. Apr 2021;34:100812.

4. Centers for Disease Control and Prevention (CDC). What to Know About HIV and COVID-19. 2021; https://www.cdc.gov/coronavirus/2019-ncov/need-extra-precautions/hiv.html]. Accessed March 1, 2021.

5. Bhaskaran K, Rentsch CT, MacKenna B, et al. HIV infection and COVID-19 death: a population-based cohort analysis of UK primary care data and linked national death registrations within the OpenSAFELY platform. Lancet HIV. Jan 2021;8(1):e24–e32.

6. Hadi YB, Naqvi SFZ, Kupec JT, Sarwari AR. Characteristics and outcomes of COVID-19 in patients with HIV: a multicentre research network study. AIDS. Nov 1 2020;34(13):F3–F8.

7. Miyashita H, Kuno T. Prognosis of coronavirus disease 2019 (COVID-19) in patients with HIV infection in New York City. HIV Med. Jan 2021;22(1):e1–e2.

8. Brown LB, Spinelli MA, Gandhi M. The interplay between HIV and COVID-19: summary of the data and responses to date. Curr Opin HIV AIDS. Jan 2021;16(1):63–73.

9. Collins LF. Persons With Human Immunodeficiency Virus and the Coronavirus Disease 2019 Pandemic: A Viral Synergy of Biology and Sociology. Clinical Infectious Diseases. 2020.

10. National Advisory Committee on Immunization (NACI). Recommendations on the use of COVID-19 vaccines [Internet]. 2021; https://www.canada.ca/en/public-health/services/immunization/national-advisory-committee-on-immunization-naci/recommendations-use-covid-19-vaccines.html]. Accessed March 1, 2021.

11. Government of Canada. Canada’s COVID-19 Immunization Plan: Saving Lives and Livelihoods [Internet]. 2020; https://www.canada.ca/content/dam/phac-aspc/documents/services/diseases/2019-novel-coronavirus-infection/canadas-reponse/canadas-covid-19-immunization-plan-en.pdf]. Accessed March 1, 2021.

12. Pebody R. Have COVID-19 vaccines been tested in people with HIV? 2021; https://www.aidsmap.com/about-hiv/have-covid-19-vaccines-been-tested-people-hiv]. Accessed May 1 2021.

13. Burns F. HIV and COVID-19 Vaccines. International Workshop on HIV and Women. Virtual. Available at https://academicmedicaleducation.com/hiv-women-2021.April 26, 2021.

14. BC Centre for Excellence in HIV/AIDS (BCCfE).CDET Committee Statement Update on the use of COVID-19 vaccines in Persons Living with HIV. May 7 2021; http://bccfe.ca/sites/default/files/uploads/publications/centredocs/covid-19_vaccine_cdet_update_final_5_pm_2021_may_9th_.pdf]. Accessed June 1, 2021.

15. MacDonald NE. Vaccine hesitancy: Definition, scope and determinants. Vaccine. Aug 14 2015;33(34):4161–4164.

16. World Health Organization (WHO). Report of the SAGE working group on vaccine hesitancy (October 2014). 2014; https://www.who.int/immunization/sage/meetings/2014/october/1_Report_WORKING_GROUP_vaccine_hesitancy_final.pdf]. Accessed March 1, 2021.

17. Bogart LM, Ojikutu BO, Tyagi K, et al. COVID-19 Related Medical Mistrust, Health Impacts, and Potential Vaccine Hesitancy Among Black Americans Living With HIV. J Acquir Immune Defic Syndr. 2021;86(2):200–207.

18. Grumbach K, Judson T, Desai M, et al. Association of Race/Ethnicity With Likeliness of COVID-19 Vaccine Uptake Among Health Workers and the General Population in the San Francisco Bay Area. JAMA Intern Med. Jul 1 2021;181(7):1008–1011.

19. Ogilvie GS, Gordon S, Smith LW, et al. Intention to receive a COVID-19 vaccine: results from a population-based survey in Canada. BMC Public Health. May 29 2021;21(1):1017.

20. Bourgeois A.C, Edmunds M, Awan A, Jonah L, Varsaneux O, Siu W. HIV in Canada—Surveillance Report, 2016. Can Commun Dis Rep. 2017;43(12):248–256.

21. Shokoohi M, Bauer GR, Kaida A, et al. Social determinants of health and self-rated health status: A comparison between women with HIV and women without HIV from the general population in Canada. PLoS One. 2019;14(3):e0213901.

22. Donaldson MA, Campbell AR, Albert AY, et al. Comorbidity and polypharmacy among women living with HIV in British Columbia. AIDS. 2019;33(15):2317–2326.

23. Kendall CE, Wong J, Taljaard M, et al. A cross-sectional, population-based study measuring comorbidity among people living with HIV in Ontario. BMC Public Health. 2014/02/13 2014;14(1):161.

24. Lourenço L, Colley G, Nosyk B, Shopin D, Montaner JS, Lima VD. High levels of heterogeneity in the HIV cascade of care across different population subgroups in British Columbia, Canada. PLoS One. 2014;9(12):e115277.

25. Shapiro GK, Tatar O, Dube E, et al. The vaccine hesitancy scale: Psychometric properties and validation. Vaccine. Jan 29 2018;36(5):660–667.

26. Ajzen I. The theory of planned behavior. Organizational Behavior and Human Decision Processes. 1991/12/01/ 1991;50(2):179–211.

27. Women’s Health Research Institute (WHRI). RESPPONSE: Rapid Evidence Study of a Provincial Population Based COhort for GeNder and SEx. 2020; https://whri.org/covid-19-respponse-study/]. Accessed February 1, 2021.

28. Loutfy M, de Pokomandy A, Kennedy VL, et al. Cohort profile: The Canadian HIV Women’s Sexual and Reproductive Health Cohort Study (CHIWOS). PloS one. 2017;12(9):e0184708–e0184708.

29. Loutfy MR, Sherr L, Sonnenberg-Schwan U, et al. Caring for women living with HIV: gaps in the evidence. Journal of the International AIDS Society. 2013;16(1):18509–18509.

30. Hankins C. Gender, sex, and HIV: how well are we addressing the imbalance? Current Opinion in HIV and AIDS. 2008;3(4):514–520.

31. Webster K, Carter A, Proulx-Boucher K, et al. Strategies for Recruiting Women Living with Human Immunodeficiency Virus in Community-Based Research: Lessons from Canada. Prog Community Health Partnersh. 2018;12(1):21–34.

32. Canadian Institutes of Health Research (CIHR). Strategy for Patient-Oriented Research - Patient Engagement Framework. 2019; https://cihr-irsc.gc.ca/e/48413.html]. Accessed March 1, 2021.

33. Kaida A, Carter A, Nicholson V, et al. Hiring, training, and supporting Peer Research Associates: Operationalizing community-based research principles within epidemiological studies by, wtih, and for women living with HIV. Harm Reduction Journal. 2019;16.

34. Harris PA, Taylor R, Thielke R, Payne J, Gonzalez N, Conde JG. Research electronic data capture (REDCap)--a metadata-driven methodology and workflow process for providing translational research informatics support. J Biomed Inform. Apr 2009;42(2):377–381.

35. Statistics Canada. Ethnic Origin Reference Guide, Census of Population, 2016. 2017; https://www12.statcan.gc.ca/census-recensement/2016/ref/guides/008/98-500-x2016008-eng.cfm]. Accessed July 1, 2020.

36. Government of British Columbia. COVID-19 List of Essential Services. 2020; https://www2.gov.bc.ca/assets/gov/family-and-social-supports/covid-19/list_of_essential_services.pdf]. Accessed July 1, 2020.

37. R Core Team. The R Project for Statistical Computing v.4.0.2. 2020; https://www.r-project.org/]. Accessed Nov 16, 2020.

38. Khubchandani J, Sharma S, Price JH, Wiblishauser MJ, Sharma M, Webb FJ. COVID-19 Vaccination Hesitancy in the United States: A Rapid National Assessment. J Community Health. Apr 2021;46(2):270–277.

39. Frater J, Ewer KJ, Ogbe A, et al. Safety and immunogenicity of the ChAdOx1 nCoV-19 (AZD1222) vaccine against SARS-CoV-2 in HIV infection: a single-arm substudy of a phase 2/3 clinical trial. Lancet HIV. Jun 18 2021.

40. Shimabukuro TT, Kim SY, Myers TR, et al. Preliminary Findings of mRNA Covid-19 Vaccine Safety in Pregnant Persons. New England Journal of Medicine. 2021;384(24):2273–2282.

41. Patterson S, Nicholson V, Milloy MJ, et al. Awareness and Understanding of HIV Non-disclosure Case Law and the Role of Healthcare Providers in Discussions About the Criminalization of HIV Non-disclosure Among Women Living with HIV in Canada. AIDS Behav. Jan 2020;24(1):95–113.

42. Kaida A, Carter A, Nicholson V, et al. Hiring, training, and supporting Peer Research Associates: Operationalizing community-based research principles within epidemiological studies by, with, and for women living with HIV. Harm Reduct J. Jul 18 2019;16(1):47.

43. Loutfy M, Greene S, Kennedy VL, et al. Establishing the Canadian HIV Women’s Sexual and Reproductive Health Cohort Study (CHIWOS): Operationalizing Community-based Research in a Large National Quantitative Study. BMC Med Res Methodol. Aug 19 2016;16(1):101.

44. Government of British Columbia. COVID-19 Immunization Plan. 2020; https://www2.gov.bc.ca//gov/content/covid-19/vaccine/plan]. Accessed June 1, 2021.

45. Government of British Columbia. British Columbia COVID-19 dashboard. 2021; https://experience.arcgis.com/experience/a6f23959a8b14bfa989e3cda29297ded]. Accessed July 8, 2021.

46. BC Centre for Disease Control (BCCDC). HIV in British Columbia: Annual Surveillance Report 2017.2019; http://www.bccdc.ca/health-professionals/data-reports/hiv-aids-reports]. Accessed June 1, 2021.

