## Supplementary Table 1 for "Intention to receive a COVID-19 vaccine by HIV status among a population-based sample of women and gender diverse individuals in British Columbia, Canada"

Supplementary Table I. Bivariable associations between vaccine hesitancy and psychological constructs and intention to receive the COVID-19 vaccine among women and gender diverse individuals in British Columbia, Canada (n=5,588)

| | Crude OR | 95% CI | P-value |
|---|---|---|---|
| WHO Lack of Vaccine Confidence Scale | 0.32 | 0.28 – 0.36 | <0.001 |
| WHO Vaccine Risks Scale | 0.52 | 0.48 – 0.56 | <0.001 |
| Attitudes toward the COVID-19 Vaccine Scale | 1.16 | 1.14 – 1.18 | <0.001 |
| Perceived Behavioural Control Scale | 1.07 | 1.05 – 1.10 | <0.001 |
| Direct Social Norms Scale | 1.27 | 1.25 – 1.30 | <0.001 |
| Indirect Social Norms: Total Scale | 1.10 | 1.09 – 1.11 | <0.001 |
| Indirect Social Norms: Family Doctor/Primary Healthcare Provider | 1.28 | 1.25 – 1.31 | <0.001 |
| Indirect Social Norms: BC Provincial Health Officer | 1.25 | 1.23 – 1.28 | <0.001 |
| Indirect Social Norms: Friends | 1.35 | 1.31 – 1.39 | <0.001 |
| Indirect Social Norms: Family | 1.31 | 1.28 – 1.34 | <0.001 |
